# Non-targeted metabolomics analysis reveals distinct metabolic profiles between positive and negative emotional tears of humans

**DOI:** 10.1101/2022.01.28.22270049

**Authors:** Hao Liang, Songye Wu, Duo Yang, Jianhua Huang, Xiaolei Yao, Jingbo Gong, Pei Liu, Lunhui Duan, Liu Yang, Qingwen Xu, Rujia Huang, Meiheriayi Maimaitituersun, Lijuan Tao, Qinghua Peng

## Abstract

**BACKGROUND:** Although the chemical components of basal, reflex, and emotional tears are different, the presence of distinctions in the tears of different emotions is still unknown. The present study aimed to address the biochemical basis behind emotional tears through non-targeted metabolomics analysis between positive and negative emotional tears of humans.

**METHODS:** Samples of reflex (C), negative (S), and positive (M) emotional tears were collected from healthy college participants. Untargeted metabolomics was performed to identify the metabolites in the different types of tears. The differentially altered metabolites were screened and assessed using univariate and multivariate analyses.

**RESULTS:** The global metabolomics signatures classified the C, S, and M emotional tears. A total of 133 significantly differential metabolites of ESI-mode were identified between negative and positive emotional tears. The top 50 differential metabolites between S and M were highly correlated. The significantly altered pathways included porphyrin & chlorophyll metabolism, bile secretion, biotin metabolism, arginine & proline metabolism and among others.

**CONCLUSION:** The metabolic profiles between reflex, positive, and negative emotional tears of humans are distinct. Secretion of positive and negative emotional tears are distinctive biological activities. Therefore, the present study provides a chemical method to detect human emotions which may become a powerful tool for diagnosis of mental disease and identification of fake tears.

## Introduction

There are three types of tear production in humans: basal, reflex, and emotional^1^. The basal and reflex tearing serve the human eyes and their physiological functions are fully understood. However, the origin and mechanism of emotional tearing has been poorly studied. Emotional crying appears to be a unique human behavior, resulting from cognitive and psychogenic brain process^2^. Although emotional tearing also originates in the eyes, it is basically useless to the organ. Emotional tears is often considered as communicative signals, such as to express the need for help or empathic responses. Further, although this mysterious behavior has fascinated both scientists and lay people alike, its research is still at ABC stage to uncover the neurophysiological and biochemical underpinnings of the behavior.

Human newborns display vocal crying when they are separated from their mothers. This behavior is consistent across different species of mammals and birds and requires no previous learning. In this period, the shedding of visible tears is triggered by strong contractions of the orbicularis oculi muscle during the production of distress vocalizations which stimulates sensitive corneal sensory nerves^3^. Real emotional tearing appears later than reflex tearing, several months after birth. According to the research conducted by Montagu^4^, it was reported that most children display emotional tearing nearly in the sixth week of life, but Vignat *et al*. found that emotional tearing occurs at about 4 months of age^2^.

Tears evolves with increasing age to serve as emotion signals that convey more complex information. Physical pain and discomfort are important triggers of tears in infants and children. However, adults and the elderly people seldom cry because of the physical condition and their weeping is usually related to different emotional states^5^. As an essential additional feature of crying in human, what is the role of emotional tears? Available, clinical literature indicates that tearful crying, in particular results in tension reduction and even has health benefits. The biochemical hypothesis suggests that the (endogenous) release of endorphins or oxytocin while crying seems catharsis to the criers in negative situations^6^. Whereas, adults cry not only in negative situations, such as losses, failures, and helplessness, but also in positive situations, such as when witnessing the intensification of relationships, prosocial behaviors, and even happiness^5^.

Tears are composed of proteins, lipids, metabolites, and electrolytes. It has already been shown that the chemical components of tears are different among basal, reflex and emotional tears ^7, 8^. However, whether the distinctions are present in tears of different emotions is still unknown. It has been found that tears of mice contains a chemosignal or pheromone. According to a study conducted by Gelstein *et al*. it was reported that human tears also contain similar chemosignal^9^. The present study hypothesized that different emotional tears (positive or negative emotions) may contain distinct substances, which would be the basis of chemosignal for tearing. Therefore, this study sought to address the biochemical basis behind emotional tears through non-targeted metabolomics analysis between the negative and positive emotional tears of humans, as well as to reveal biomarkers of the emotions through bioinformatic identification.

## Methods

### Study population

#### Ethics statement

The protocol of the current study was approved by the ethics committee of Jili Hospital and was conducted in accordance with the Decoration of Helsinki. The study was registered in the China Clinical Trial Registration Center (Registration No. ChiCTR2100047025, http://www.chictr.org.cn/showproj.aspx?proj=127637). Moreover, written informed consent was obtained from each participant.

#### Inclusion and exclusion criteria for the participants

The inclusion criteria for the study participants were:

- Age between 17 and 35 years.
- No gender limit.
- Physical and mental health.

The exclusion criteria were as follows:

- A refractive measurement of <-6.0 D or > + 5.0 D.
- Catarrhal inflammation in recent 7 days due to upper respiratory tract infection and conjunctivitis among others.
- Eye surgery in recent 6 months.
- Febrile illnesses in recent 7 days.
- Mental illness history such as depression, autism, and schizophrenia or recent psychic trauma.
- Long-time (>1 month) insomnia.
- Unhealthy habits such as smoking and drinking.

All the volunteers received ophthalmic examinations between June 1st and September 30th of 2021. The examination included visual acuity, refraction, slit-lamp, and tear film breakup time to assure the healthy state of their ocular surface. Fifty healthy students (30 females and 20 males) from Hunan University of Chinese Medicine met the inclusion criteria and were finally recruited into the current study.

### Study design and sample collection

#### Tears types and identification

Essential balm was applied on the bags beneath eyes to stimulate the reflex tears. Negative movies (themes of sadness, losing, betrayal, torture, persecution, and discrimination) were selected to help produce tears of negative emotions such as distressed, upset, guilty, scared, nervous, and afraid. On the other hand, positive movies (themes of happiness, love, friendship, faith, and loyalty) were selected to help produce tears of positive emotions such as excited, strong, enthusiastic, inspired, and determined.

By watching a lot of the selected movies, the research team of the current study voted for the movies that could induce negative or positive emotional tears as follows:

#### Negative

- The Chinese medical documentary *人 间 世 (Life Matters)* [S2EP01, https://www.youtube.com/watch?v=knJ3t-GeUyc]. It’s a story documenting the end stage of children with cancers in hospital.
- The movie *Manchester by the Sea*.

#### Positive

- The movie *Hachi: A Dog’s Tale (2009)*
- The movie *The Pursuit of Happyness*

Reflex tears from the participants induced by essential balm were labelled as C samples (control). The emotional tears were induced by watching different movies. When the emotional tears were successfully induced and collected, the participants were immediately required to finish the Positive and Negative Affect Schedule (PANAS-SF) which is a scale that consists of different words that describe feelings and emotions^10^. The participants described the emotions according to the scale at that moment to shed tears. If the positive affect score of PANAS-SF range from 10 to 50, the tears sample was labelled as M (positive, induced by moving scenes), or else the sample is classified as S (negative, induced by sad scenes) when the negative affect score ranged from 10 to 50. To avoid individual variations, the three types of tears were all from the same participants.

#### Tears collection

Each participant was requested to watch movie and collect samples in a private room at between 16:00 and 21:00, and the sleeping time must exceed 6 hours during the past 24 hours. Further, the contact lenses were also forbidden to use in the past 24 hours. The face was washed to clean any make-up on eyelid, eyelash, and face before the sample collection process.

Schirmer tear test strips were used for collecting tears from both eyes. The researchers bend a Schirmer strip at the preformed notch at 90° and placed into conjunctival sac of the participant at the junction of the mid and temporal thirds of the lower lid when first drop of tears flow out of the eyelid. A 35 mm Schirmer strip is equal to 35 μL volume, the length of the moistened area was measured using the millimeter scale on the strip. Three Schirmer strips (wetting of more than 25 mm of each strip) collecting each type of tears from every subject were placed in a 2.0 mL sterile cryogenic vial, frozen, and then immediately transferred to –80°C refrigerator until metabolomic analysis was carried out. The collecting interval between different types of tears in each subject was more than 24 hours. Finally, different types of tears (reflex, positive and negative emotional tears) were completely collected from the 12 participants. The workflow of this study was shown in **Figure 1**.

**Figure 1.**
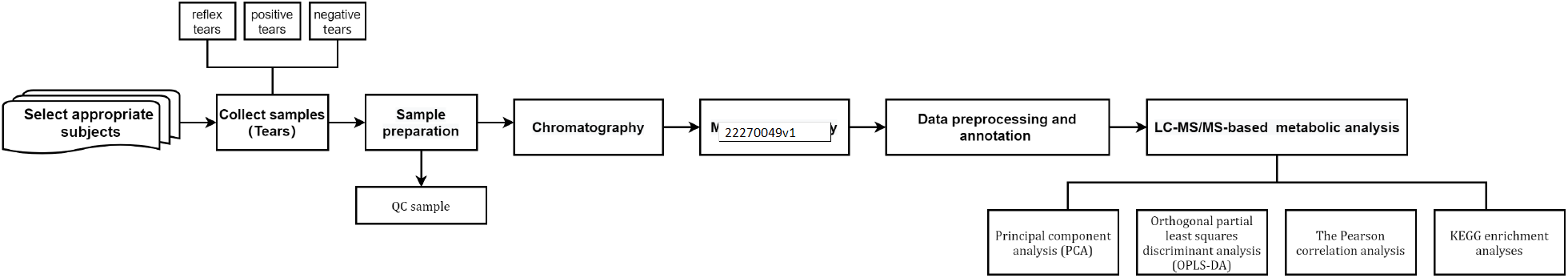
Workflow of tears collection and analysis

### Metabolomics analysis

#### Sample preparation

The sample were taken out from the -80^°^Crefrigerator and thawed on ice. The redundant part was cut off to keep each of Schirmer strips at 24 mm length and 70% methanol water internal standard extractant (300 μL) was mixed in corresponding Eppendorf tube with sample number, vortexed for 5 minutes, sonicated in ice water bath for 10 minutes, and then allowed to stand still at -20°C for 30 minutes. Next, the mixture was centrifuged (12000 rpm, 4°C) for 10 minutes, and 200 μL of the supernatant was transferred to a new centrifugal tube. Finally, this was centrifugated (12000 rpm, 4°C) for 3 minutes and 150 μL of supernatant was taken for LC-MS/MS analysis. Moreover, 20 μL of each tears sample from each group was pipetted into a centrifuge tube to prepare a QC sample for LC-MS/MS analysis.

#### Chromatography

Chromatographic separation was performed using a Shimadzu LC20AD UPLC system (Shimadzu Scientific Instruments, Kyoto, Japan) which was equipped with a Waters ACQUITY UPLC HSS T3 C18 column (1.8 µm, 2.1 mm*100 mm). The column temperature was set at 40°C and at a flow rate of 0.40 mL/min. The gradient system consisted of 0.1% formic acid in ultrapure Water in mobile phase A and 0.1% formic acid in acetonitrile in mobile phase B (0 minutes, 5% B; 11 minutes, 90% B; 12 minutes, 90% B; 12.1 minutes, 5% B; 14 minutes, 5% B).

#### Mass spectrometry

Tandem mass spectrometry (MS/MS) was performed using a AB Sciex TripleTOF® 6600 quadrupole time-of-flight (QTOF) mass analyzer (AB Sciex LLC, MA, USA). Mass calibration was conducted daily according to recommended procedure provided by the manufacturer. An electrospray ionization source (ESI source) was used for mass spectrometric detection in the positive and negative ionization mode. The QC samples were used for MS/MS data acquisition. The source parameters were set as shown in S_Table 1.

### Data analysis

#### Data preprocessing and annotation

The original data file obtained through LC-MS analysis was first converted into mzML format by ProteoWizard software. Peak extraction, alignment, and retention time correction were performed using XCMS program. Then, the support vector regression (SVR) method was used to correct the peak area^11^. The peaks with deletion rate > 50% were filtered in the samples of each group. In addition, the metabolic identification information was then obtained by searching the self-built database of the lab and integrating the public databases (Metlin & HMDB) as well as metDNA.

#### Statistical analysis and visualization

Principal component analysis (PCA) was first used to reduce the dimensionality of the multidimensional dataset. Orthogonal partial least squares discriminant analysis (OPLS-DA) was then used to explore the main metabolic distinction between the different types of tears (Q2>0.40). Validation plot was employed to assess the validity of OPLS-DA model by comparing 200 random permutations of the Y variable and the goodness of fit (R2Y and Q2)12. The corresponding variable importance in the projection (VIP values) was calculated in OPLS-DA model and VIP value > 1 indicated a significant difference. The T-test (P value < 0.05) and fold change (FC, FC ≥ 2 or FC ≤ 0.5) were also applied to screen the potential differential metabolites. The Pearson correlation and cluster analyses were conducted to determine the correlation between the differentially altered metabolites. Further, the pathway functions were used to investigate the differentially altered metabolites. Pathway and KEGG enrichment analyses were applied to identify the related pathways to emotional tears. All of the statistical analyses and visualization were performed on the R platform (version 4.1, The R Foundation, Vienna, Austria) software.

## Results

### Demographic information

Thirty-six tear samples (12 samples of each type of tears - reflex, positive, and negative emotional) were successfully collected from the 12 participants (11 females and 1 male). No abnormal signs were found from the ophthalmic examinations in the subjects. The details of ocular surface examination were as shown in S_Table 2.

### Metabolomic analysis

#### QC samples analysis

In the ESI+ and ESI-mode, the total ion chromatogram (TIC) of the tears in the QC samples was as shown in S_Figure 1. The overlap of the TIC curves was high, exhibiting good stability of mass spectrometry for the same sample at different times. The PCA plot of all the samples showed that the QC samples were tightly clustered (S_Figure 2) which indicated a good analytical reproducibility of the current metabolomics study.

#### Metabolic profiles of different types of tears

The PCA plot displayed a tendency of distinction in metabolic profiles between different types of tears (reflex, positive, and negative emotional). The OPLS-DA plot (Figure 2) revealed a remarkable separation in reflex tears (C) vs. negative emotional tears (S) and reflex tears (C) vs. positive emotional tears (M) of ESI+ mode, as well as in C vs. M and S vs. M of ESI-mode. Specifically, the OPLS-DA score between C and S resulted in a R2Y = 0.996 and Q2 = 0.609 for ESI+ mode, as well as a R2Y = 0.692 and Q2 = 0.184 for ESI-mode. The OPLS-DA score between C and M showed a R2Y = 0.992 and Q2 = 0.616 for ESI+ mode, as well as a R2Y = 0.999 and Q2 = 0.987 for ESI-mode. The OPLS-DA score between S and M resulted in a R2Y = 0.986 and Q2 = 0.153 for ESI+ mode, as well as a R2Y = 0.952 and Q2 = 0.491 for ESI-mode. Moreover, the results of the validation analysis as presented in S_Figure 3 supports the reliability and good fitting of the OPLS-DA model in C vs. S and C vs. M of ESI+ mode, as well as C vs. M and S vs. M of ESI-mode, because the p values of permuted R2Y and Q2 were all under 0.05.

**Figure 2.**
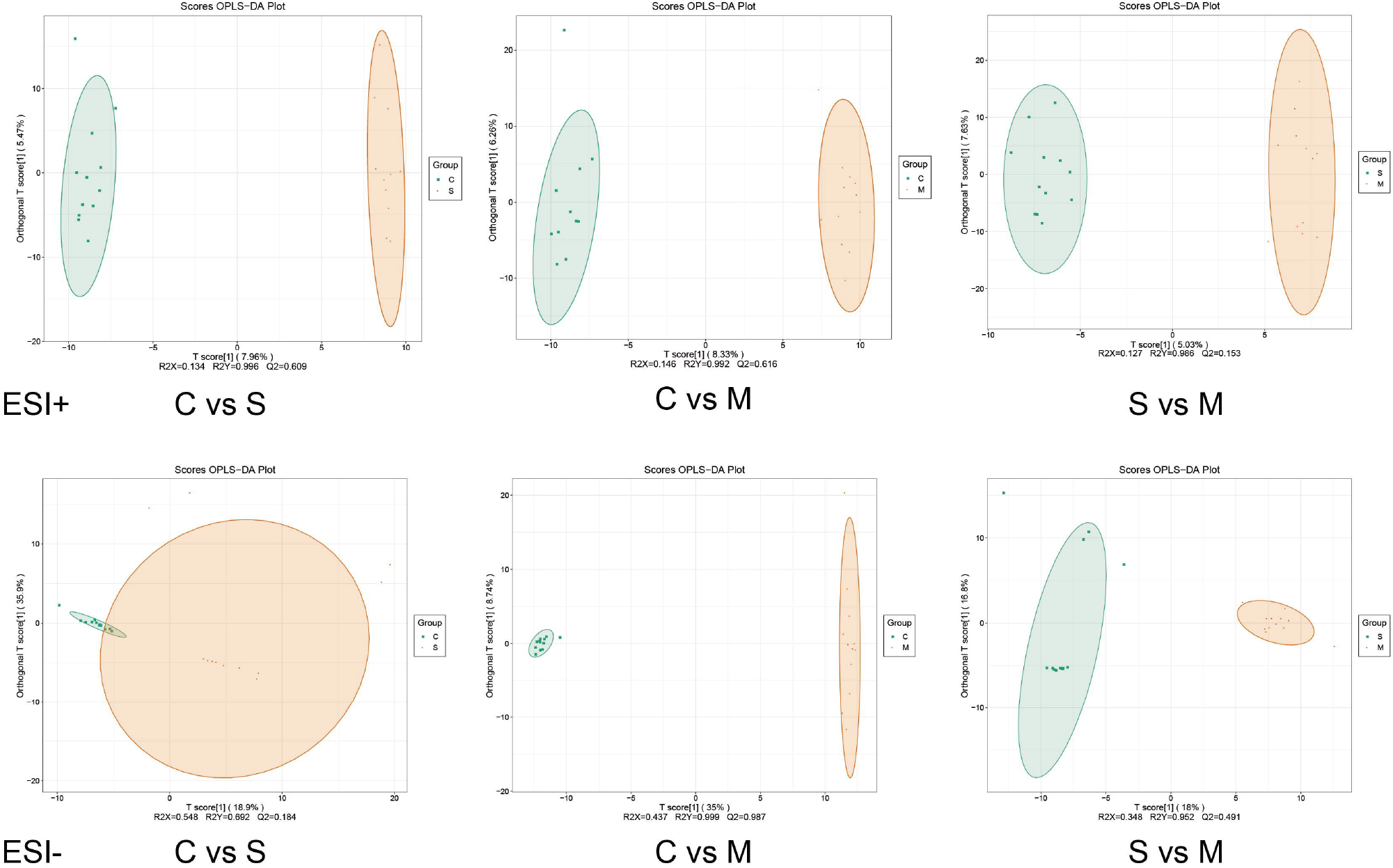
OPLS-DA score plots of the multivariate statistical analysis between reflex tears, negative emotional tears and positive emotional tears. C Group: reflex tears; S Group: negative emotional tears; M Group: positive emotional tears.

The potential differential metabolites were screened based on the screening criteria (VIP value > 1, P value < 0.05, and FC ≥ 2 or FC ≤ 0.5). There were 90 and 80 differential metabolic molecules in C vs. S and C vs. M of ESI+ mode respectively. The top five influential metabolites (VIP value) between C and S were *γ*-Dodecalactone, 4-Epi-Brassinolide, PA(18:3(6Z,9Z,12Z)/16:0), 12-Ketodeoxycholic acid, and 2-(2,6-Dimethoxy-4-prop-2-enylphenoxy)-1-(3,4,5-trimethoxyphenyl)propan-1-ol. Further, the top five influential metabolites between C and M were 2-Phenylacetamide, (S)-Cotinine N-oxide, *γ*-Dodecalactone, Lauric acid, Brinzolamide. Moreover, 158 and 133 differential metabolites were identified in C vs. M and S vs. M of ESI-modes, respectively. The top five influential metabolites between C and M of ESI-mode were Alprazolam, Dethiobiotin, (-)-threo-Iso(homo)2-citrate, 1,3-Diacetoxy-4,6,12-tetradecatriene-8,10-diyne and Maculine. While, the top five influential metabolites between S and M were 1,3-Diacetoxy-4,6,12-tetradecatriene-8,10-diyne, Indole-3-acetic acid FFA (18:0), Nopaline, and N6-Acetyl-L-Lysine. The overall influential metabolites in the current study were displayed as the volcano plots (S_Figure 4) and Venn plots (Figure 3). A total of 133 significantly differential metabolites of ESI-mode were identified between negative and positive emotional tears. The detailed information of tears metabolites was as shown in S_Table 3 (https://github.com/hao203/tears/blob/main/differential/S_vs_M_neg_filter_identi_known.xlsx).

**Figure 3.**
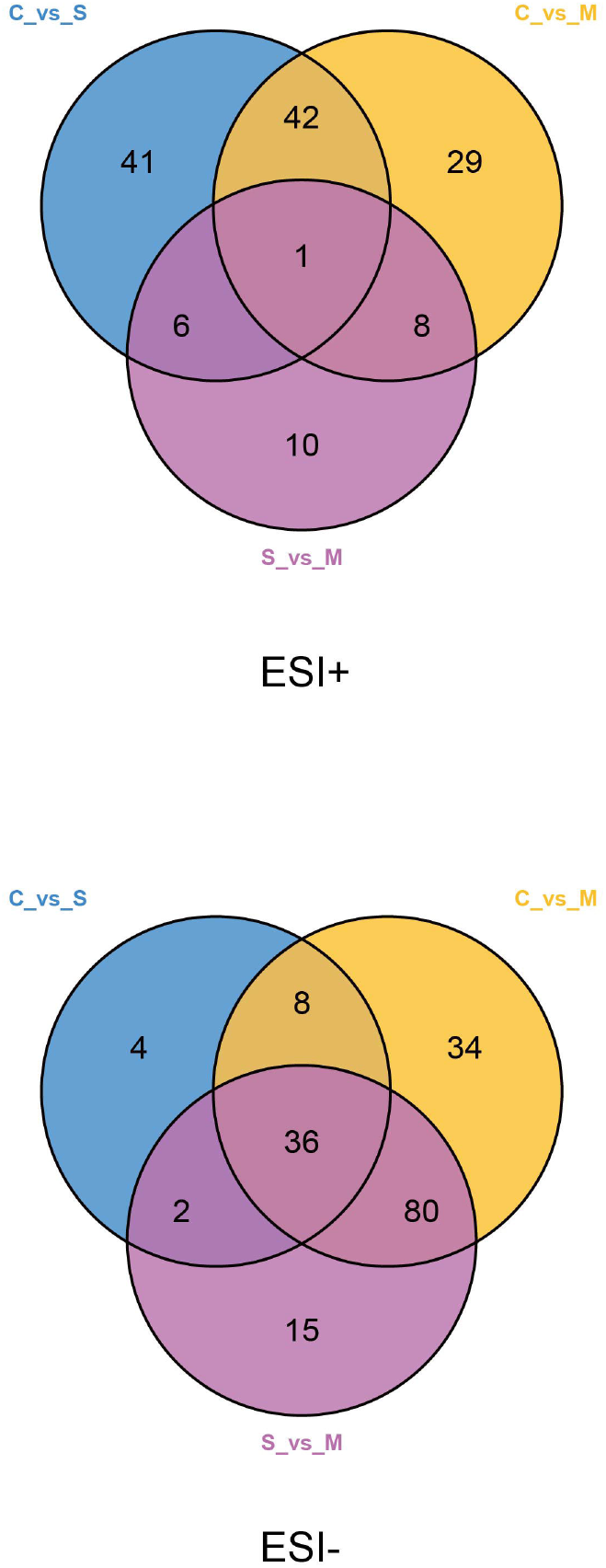
Venn plots of the screened metabolites between reflex tears, negative emotional tears and positive emotional tears.

#### Correlation analysis of the differential metabolites

The relationship between the differential metabolites associated with different classifications of tears was accessed through cluster and Pearson correlation analyses. The heatmaps of cluster analysis showed the relative intensity distribution and relationship of these metabolites. The metabolites in different groups were clearly divided using color and the downregulated and upregulated metabolites were closely clustered (S_Figure 5). Furthermore, the Pearson correlation analysis was performed to explore the relations of the top 50 differential metabolites (VIP value) in each mode. The heatmaps of correlation showed that the metabolites in C vs. M and S vs. M of ESI-mode were highly correlated (Figure 4).

**Figure 4.**
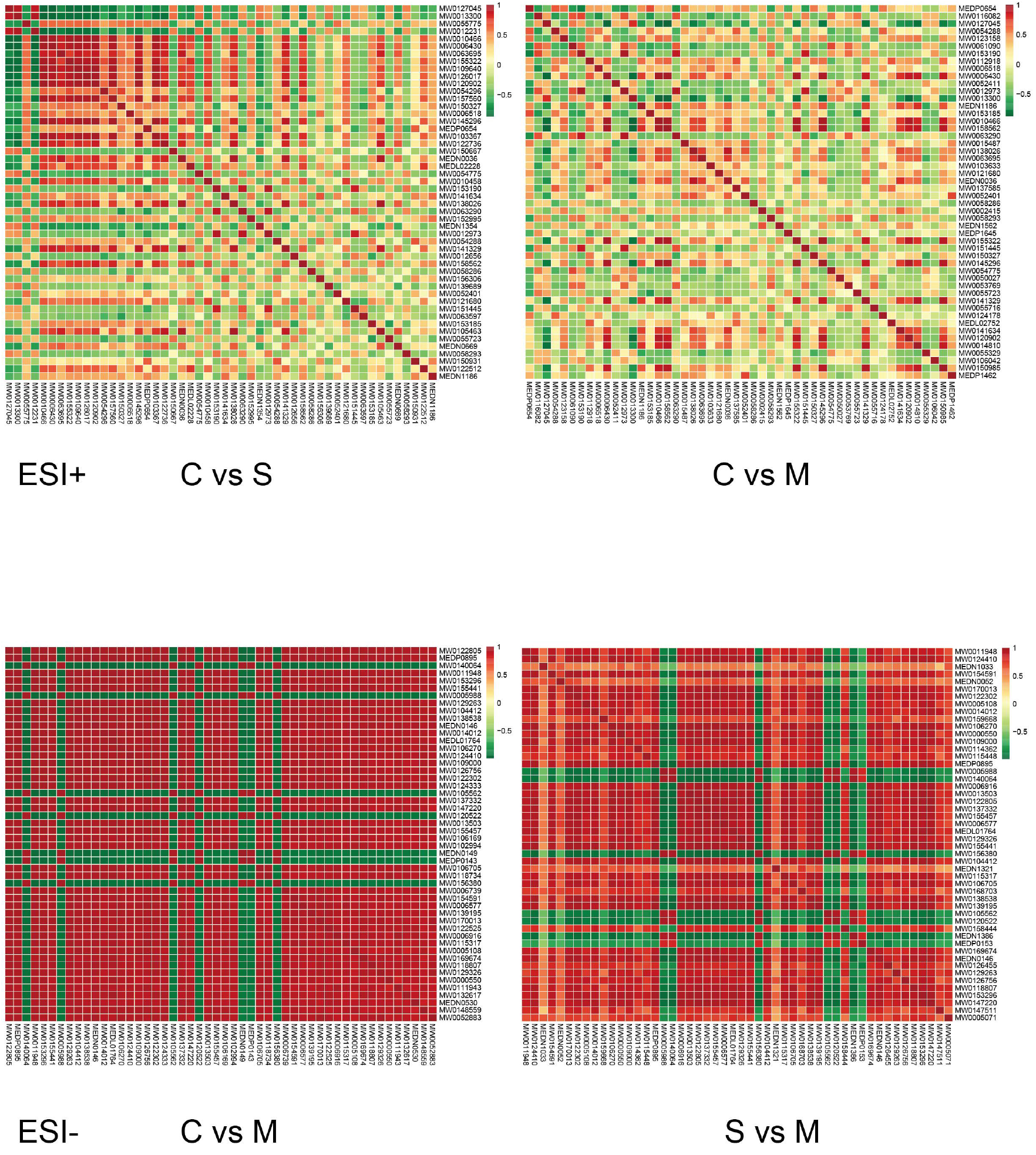
Correlation plots of the top 50 identified metabolites between reflex tears, negative emotional tears and positive emotional tears. C Group: reflex tears; S Group: negative emotional tears; M Group: positive emotional tears.

#### Analysis of the metabolic pathways

ClusterProfiler package was used to analyze the relationship between the differentially altered metabolites and all the annotated metabolites were mapped into biochemical pathways to reveal a mechanistic interpretation. The KEGG enrichment scatter plot was used to show the significantly related pathways (p<0.30 and Rich Factor>0.20). The main affected pathways of C vs. S (Figure 5) were arachidonic acid metabolism, serotonergic synapse, and estrogen signaling pathway, Gonadotropin-releasing hormone (GnRH) secretion, and beta−Alanine secretion and metabolism. The main pathways involved in C vs. M (Figure 5) are propanoate metabolism, mineral absorption, phenylalanine metabolism, and nitrogen metabolism of ESI+ mode, and biotin metabolism and caffeine metabolism of ESI-mode. Meanwhile, the prominently related pathways of S vs. M (Figure 5) were porphyrin & chlorophyll metabolism, bile secretion, biotin metabolism, arginine & proline metabolism, and phosphonate & phosphinate metabolism.

**Figure 5.**
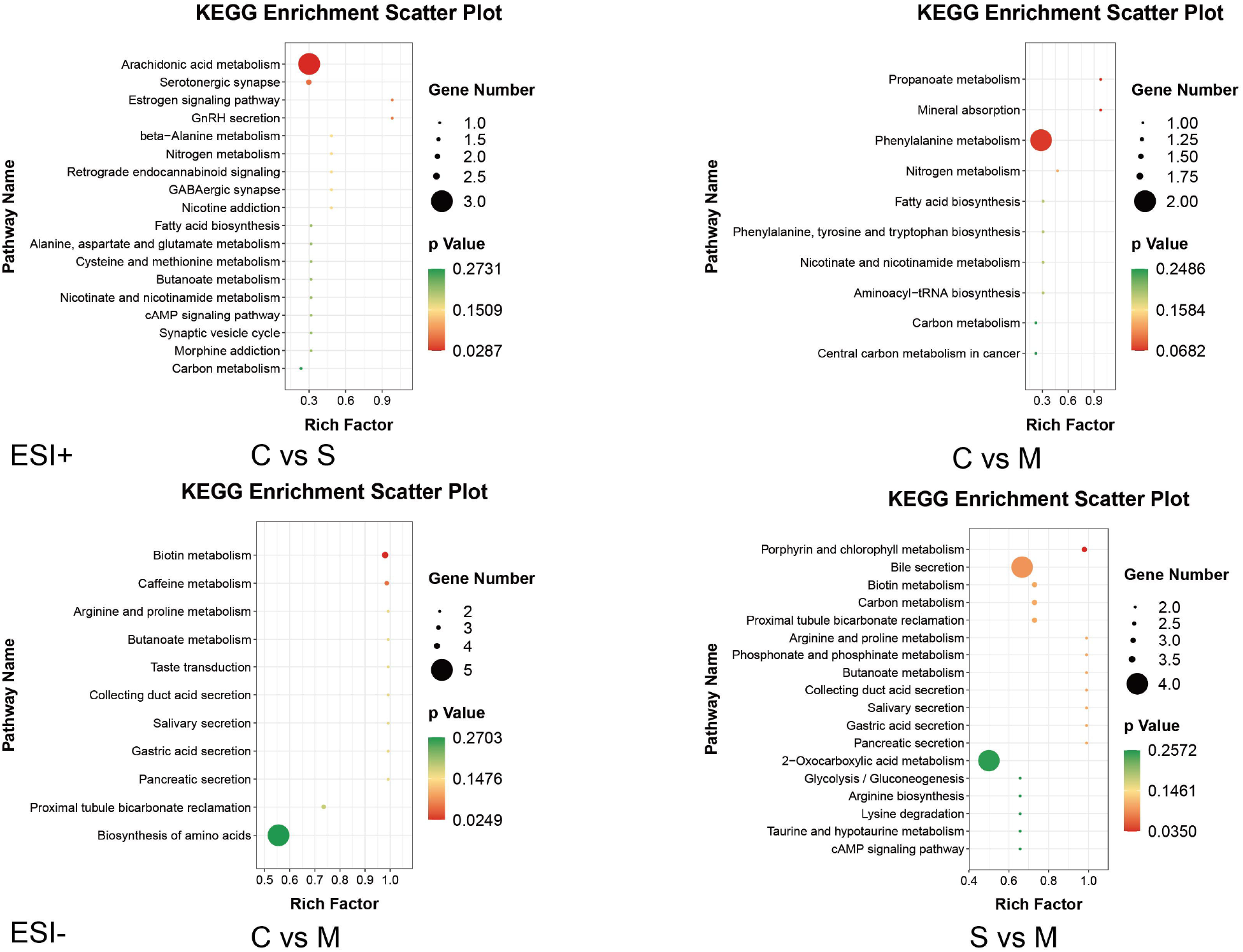
KEGG enrichment scatter plots of the identified metabolites between reflex tears, negative emotional tears and positive emotional tears. C Group: reflex tears; S Group: negative emotional tears; M Group: positive emotional tears.

## Discussion

An untargeted metabolomics study based on LC-MS/MS technique was performed in the present study to investigate differential metabolic signatures in different types of tears. The comparison of metabolite features between different types of tears has shown differing profiles. Visual separation is explicit from OPLS-DA score plots and the Q2 values in C vs. S and C vs. M of ESI+ mode, as well as C vs. M and S vs. M of ESI-mode are all greater than 0.40, which is acceptable for classification in human metabolomics studies^13, 14^. Specifically, in the current study, a total of 133 differential metabolites were identified between positive and negative emotional tears in ESI-mode. Further, the correlation analysis showed a significantly positive or negative correlation between the top 50 differential metabolites based on VIP score. Moreover, it was evident that some metabolites were only up-regulated in positive emotional tears, such as Indole-3-acetic acid (MW0124410) and free fatty acids 18:0 (FFA 18:0, MEDN1033). While some metabolites were up-regulated in both positive and negative emotional tears, the levels of the metabolites were higher in positive emotional tears, such as 1,3-Diacetoxy-4,6,12-tetradecatriene-8,10-diyne (MW0011948) and Nopaline (MW0154591). Besides, some metabolites were down-regulated in positive emotional tears, such as (-) -threo-Iso (homo) 2-citrate (MW0140064) and Aloe-emodin (MW0005988). The results of the current study indicated that metabolites are significantly different between positive and negative emotional tears. Therefore, the filtered metabolites are potential biomarkers for detection of real emotions in humans.

Tears are made up of water, electrolytes, proteins, lipids, and mucins that form layers on the surface of eyes^15^. It has been reported that the compositions significantly vary in different types of tears (basal, reflex, and emotional)^16^. Humans are the only mammals known to produce tears in response to an emotional state, such as out of joy or grief. As part of human emotional package, secretion of emotional tears may also serve a biological function by excreting stress-inducing hormones built up through times of emotional distress^17^, and a form of social signaling, such as eliciting help and support from those around you. Emotional tears contains a higher protein content. Therefore, it’s more viscous, sticking to the skin, and taking longer to roll down the face^18^. On the contrary, reflex tears are waterier to help you wash out any irritations to your eyes from foreign particles or vapors. It contains more of antimicrobial compounds known as lysozyme and defensin peptides to prevent infections^19^.The results of the current study also revealed that the levels of some anti-bacterial metabolites, such as *γ*-Dodecalactone, are higher in reflex tears than in the other types of tears^20^.

Analysis of the metabolic pathways in the present study explored the mechanism behind the production of different emotional tears. The KEGG enrichment of C vs. S has revealed that the production of negative emotional tears is related to regulations of some synapses in brain such as serotonergic synapse and GABAergic synapse, and regulations of a series of endocrine hormones, such as estrogen signaling pathway and GnRH secretion. Importantly, it also involves arachidonic acid metabolism, which plays a key role in inflammation^21, 22^. Therefore, this indicates that the process of negative emotional crying may induce fluctuation of sex hormones and inflammatory activities.

On the other hand, the KEGG enrichment of C vs. M in the current study showed that the shedding of positive emotional tears is closely correlated to biotin metabolism and caffeine metabolism. Brain areas demanding a higher metabolism of biotin such as the centers of the auditory and visual activity^23^, shows that biotin plays a pivotal role in activating carboxylases and development of neurological diseases^24^. Caffeine as the most commonly used stimulant drug for brain activity, it improves vigilance and cognition^25^ and the engendering positive emotional tears may be similar to caffeine stimulation.

Besides, positive emotional tears shedding is relevant to arginine and proline metabolism. Previous studies have identified the panel of metabolites associated with depression in animal models and patients^26, 27^. The levels of arginine, proline, taurine, glycine, and alanine are higher in depressed individuals than in healthy ones. Microbial functions and metabolites such as proline, converging into glutamate/GABA metabolism are linked to depression. For instance, proline supplementation in mice exacerbated depression along with microbial translocation^28^. Additionally, positive emotional tears production is related to a series of external secretions pathways, such as salivary secretion, gastric acid secretion and pancreatic secretion. It’s consistent with previous report that the activity of brain electrophysiological and salivary secretion can improve negative emotions of people and reduce amino acids (arginine, proline, histidine, and taurine) concentrations in saliva^26^. In summary, it is evident that secretion of either positive or negative emotional tears involves different biological activities.

Psycho-emotional weeping demonstrates not only mental responses to the environment, but also elicit sympathy and social support from observers^29, 30^. Fake tears or “crocodile tears” are an insincere tearing display that can sometimes be used to manipulate and deceive, for instance, the disguise to get off charges in court^31^. They originate from the ancient Greeks who had an anecdote in which crocodiles would pretend to weep while luring their prey in. “Crocodile tears” are typically related to the simulated tears of celebrities and politicians, and conveying fabricated remorse during criminal court proceedings^31, 32^. The accurate detection of emotional deception is crucial in the social credit system, such as in commercial negotiation, charitable causes, and court trials. The present research has provided a convenient and promising way to expose emotional deception by chemical approach.

The main limitation of this study is the small sample size. Although,50 subjects were recruited in the current study, only 12 of them (24%) successfully completed the emotional tears collection and this could because for an adult person to cry in front of a collector is rather embarrassing and difficult. Thus, the diagnostic performance of the metabolites cannot be evaluated by receiver operating characteristic (ROC) curve and area under the curve (AUC) because of the limited sample size. However, the results are still convincible, because in contrast to other metabolomics studies, the samples of different types in the current study were from the same 12 subjects, without any individual variations. Another limitation was the gender issue for crying, whereby women were definitely better at it than men and hence only one man (1/20) has finished the emotional tears collection. Furthermore, there is no tears metabolite databases, and the KEGG enrichment in the present study was simply based on the traditional algorithms and metabolite databases of blood and urine. Therefore, results of metabolic pathways analysis in the present study may be inconsistent with the reality. A tears omics database should be established in the future for biomarkers exploration and enrichment analysis.

## Conclusion

In conclusion, the present study analyzed the metabolites of 36 samples of tears from 12 participants to investigate the metabolic characteristics between reflex, positive, and negative emotional tears using non-targeted LC-MS/MS metabolomics. It was evident that there are dramatic alterations of the metabolites in the three types of tears which suggest that tears metabolomics may possess great potential for human emotional detection. Moreover, secretion of positive and negative emotional tears is shown to be different biological activities through analysis of metabolic pathways mainly involving biotin metabolism, arginine, and proline metabolism among others. Therefore, the findings of the current study contribute a chemical method for detecting human emotions and it may become a powerful tool for diagnosis of mental disease and identification of fake tears.

## Data Availability

Raw and processed datasets of mass spectrum generated during and/or analyzed during the current study are available in the China National GeneBank DataBase (CNGBdb) Repository, https://db.cngb.org/search/metabolize/METM0000028/. Furthermore, the datasets of all the identified metabolites information can be downloaded from https://github.com/hao203/tears

https://db.cngb.org/search/metabolize/METM0000028/

## Conflict of interest

The authors declare that they have no conflict of interest.

## Acknowledgments

We wish to thank the staff of Genecreate Biological Engineering Co., Ltd. for the technical assistance in MS analysis of the samples in the present study. We are also grateful to the participants in the tears collection for their invaluable contribution.

## Funding

This research study was supported by the grants of China Postdoctoral Science Foundation (No. 2020M682578), Science and Technology Innovation Program of Hunan Province (No.2020RC2061), Guidance Project of Liu Liang Academician Workstation (21YS002), and Key Projects of TCM Scientific Research Program in Hunan Province (No.201901).

## Supplementary files

All the supplementary files are available from Github Repository, https://github.com/hao203/tears/tree/main/files

**S_Figure 1** Total ion chromatogram (TIC) of the tears in the QC samples.

**S_Figure 2** Principal component analysis of the whole samples. C Group: reflex tears; S Group: negative emotional tears; M Group: positive emotional tears.

**S_Figure 3** Validation analysis by comparing the goodness of fit (R2 and Q2) of the PLS-DA models with the goodness of fit of 200 Y-permutated models. C Group: reflex tears; S Group: negative emotional tears; M Group: positive emotional tears.

**S_Figure 4** Volcano plots of the overall influential metabolites. C Group: reflex tears; S Group: negative emotional tears; M Group: positive emotional tears.

**S_Figure 5** Heatmaps of cluster analysis of the overall influential metabolites. A: ESI+ C vs. S; B: ESI+ C vs. M; C: ESI– C vs. M; D: ESI– S vs. M; C Group: reflex tears; S Group: negative emotional tears; M Group: positive emotional tears.

**S_Table 1** Source MS/MS parameters

**S_Table 2** Ocular surface examinations of the participants

**S_Table 3** Significantly differential metabolites of ESI-mode between negative and positive emotional tears.

## Notes

### Competing Interest Statement

The authors have declared no competing interest.

### Clinical Trial

ChiCTR2100047025

### Author Declarations

The protocol of the current study was approved by the ethics committee of Jili Hospital and was conducted in accordance with the Decoration of Helsinki.

